# The efficacy of DNA MMR enzyme immunohistochemistry as a screening test for hypermutated gliomas

**DOI:** 10.1101/19012005

**Authors:** Matthew McCord, Alicia Steffens, Rodrigo Javier, Kwok-Ling Kam, Kathleen McCortney, Craig Horbinski

## Abstract

A subset of gliomas has DNA repair defects that lead to hypermutated genomes. While such tumors are resistant to alkylating chemotherapies, they may also express more mutant neoantigens on their cell surfaces, and thus be more responsive to immunotherapies. A fast, inexpensive method of screening for hypermutated gliomas would therefore be of great clinical value. Since immunohistochemistry (IHC) for the DNA mismatch repair (MMR) proteins Msh2, Msh6, Mlh1, and Pms2 is already used to screen for hypermutated colorectal cancers, we sought to determine whether that panel might have similar utility in gliomas. MMR IHC was scored in 100 WHO grade I-IV gliomas with known tumor mutation burdens (TMB), while blinded to TMB data. Eight of 100 cases showed loss of one or more MMR proteins by IHC, and all 8 were hypermutated. Among the remaining 94 gliomas with intact MMR IHC, only one was hypermutated; that tumor had an inactivating mutation in another DNA repair gene, *ATM*. Overall accuracy, sensitivity, and specificity were 99%, 89%, and 100%, respectively. The strongest correlates with hypermutation were prior TMZ treatment, *MGMT* promoter methylation, and *IDH1* mutation. Among MMR-deficient hypermutated gliomas, 50% contained both MMR-lost and MMR-retained tumor cells. Together, these data suggest that MMR IHC could be a viable front-line screening test for gliomas in which immunotherapy is being considered. They also suggest that not all cells in a hypermutated glioma may actually be MMR-deficient, a finding that might need to be considered when treating such tumors with immunotherapies.

## INTRODUCTION

Gliomas are the most common primary brain tumors in adults [19]. Standard of care includes surgical resection followed by adjuvant radiation and temozolomide (TMZ), a DNA alkylating agent [24]. However, tumors nearly always recur and lose sensitivity to adjuvant therapy.

When GBMs are challenged with TMZ, recurrent subclones often emerge with inactivating mutations in genes encoding DNA mismatch repair (MMR) enzymes, most commonly *MSH2, MSH6, MLH1*, and *PMS2* [7, 11, 28]. Glioma tumor mutation burden (TMB) is normally ∼1 mutation per megabase (Mb) of DNA [2], but MMR defects can lead to a “hypermutator” phenotype, defined as TMB >20 mutations/Mb DNA [10, 11, 14]. Hypermutated tumor cells tend to display more mutant proteins on their surfaces, making them potentially more vulnerable to immunotherapies like PD-1 and PD-L1 checkpoint inhibitors [4, 9]. Other forms of hypermutated cancers have shown promising responses to immune checkpoint inhibitors [20], and there is great interest in this strategy for hypermutated gliomas [6, 10].

Next generation sequencing (NGS) is the current gold standard for detecting DNA MMR defects and quantifying TMB, but larger panels are costly and have prolonged turnaround time. Targeted NGS panels, such as Glioseq [18], are less expensive and faster, but usually do not screen for MMR mutations and do not cover enough of the genome to reliably determine TMB.

A standardized quartet of IHC stains (Msh6, Msh2, Mlh1, and Pms2) is used to detect loss of normal MMR gene expression in colorectal cancers [22]. Because most pathology laboratories already have this MMR IHC panel for routine use, we sought to determine its utility as a screening test for hypermutated gliomas.

## METHODS

The cohort consisted of 100 World Health Organization (WHO) grade I-IV gliomas from the Northwestern Nervous System Tumor Bank with known TMB and MMR gene mutations, as determined by Tempus xT NGS covering approximately 600 genes (**Table 1**). Case collection was done under a Northwestern Institutional Review Board-approved protocol. IHC was performed using 4 different primary antibodies, including Msh2 (Cell Marque G219-1129, 1:700), Msh6 (Cell Marque 44 (287M-15), 1:100), Mlh1 (Leica NCL-L-MLH1, 1:100), and Pms2 (Cell Marque MRQ-28 (288M-15), 1:50). Formalin-fixed, paraffin-embedded 4 µm thick tissue sections were baked at 60°C for 30-60 minutes before being deparaffinized and re-hydrated. Antigen retrieval for Msh6, Msh2, and Pms2 was achieved using a Universal Retrieval (Abcam) buffer in a decloaking chamber reaching 110°C for 5-20 minutes. Antigen retrieval for Mlh1 used a citrate buffer (pH 6) in a decloaking chamber reaching 110°C for 10 minutes. Slides were cooled to room temperature and washed in TBS before neutralizing endogenous peroxidase (Biocare Peroxidase 1). Slides were then treated with a serum-free casein background block (Biocare Background Sniper) before pre-incubation in a 10% goat serum block for 60 minutes. Primary antibody was added to the slides for overnight incubation at 4°C. After incubation, slides were washed in 3 5-minute washes with TBS-T before incubating in HRP polymer (Biocare MACH 4 Universal HRP Polymer). Reaction products were visualized with DAB (Biocare Betazoid DAB Chromogen Kit). Slides were counterstained with hematoxylin, dehydrated, and mounted with xylene-based mounting media.

**Table 1:**
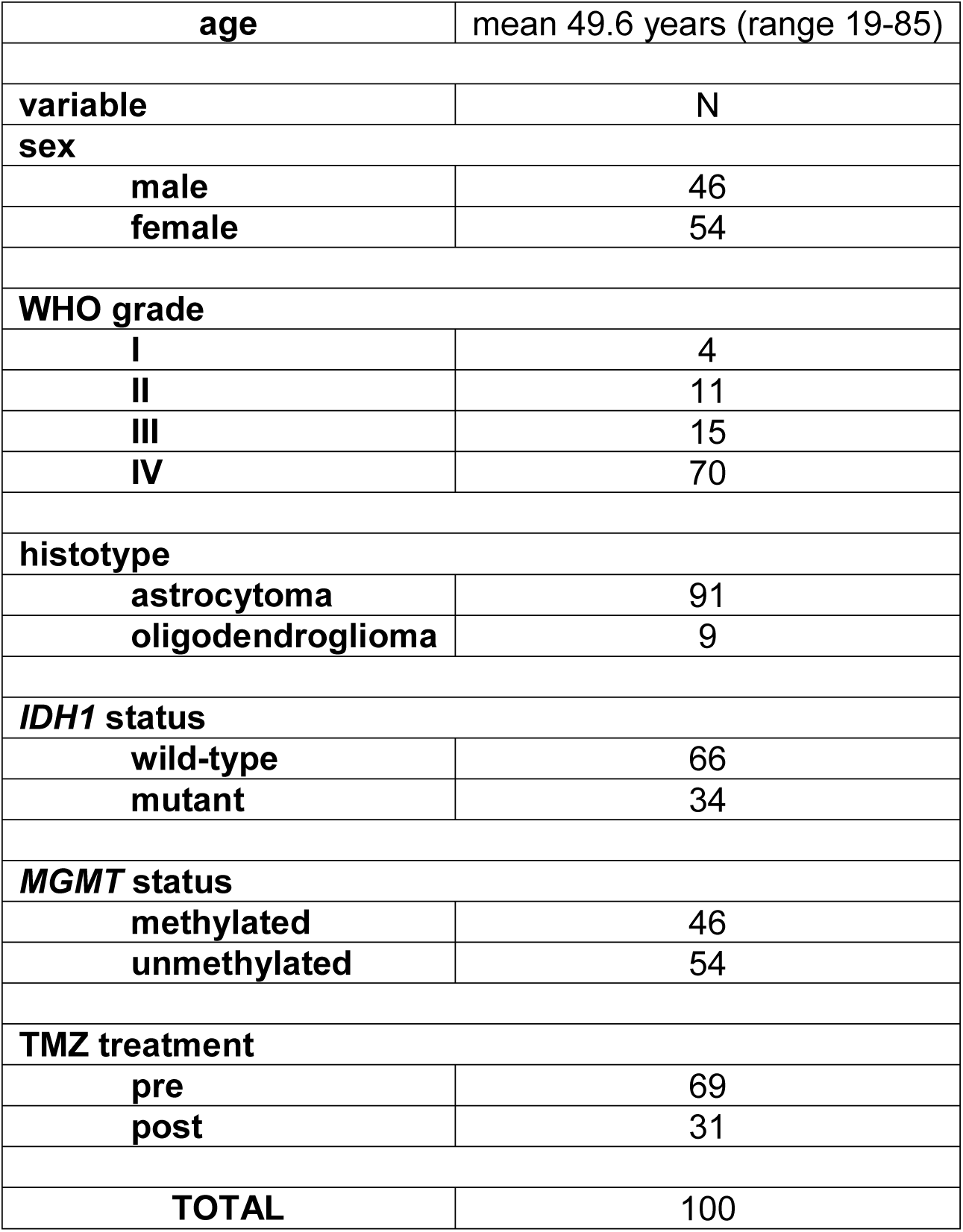
Cohort characteristics. Since cohort N=100, numbers are the same as percentages. TMZ=temozolomide.

Each IHC marker was examined under light microscopy by two independent reviewers (MM and CH) while blinded to NGS data and TMB. Each IHC marker was scored as “retained” or “lost.” In cases with lost MMR expression, the pattern (homogeneous versus heterogeneous) was noted. Nonneoplastic cells within each glioma (e.g. lymphocytes, endothelial cells) served as internal positive controls. Interobserver discrepancies were resolved by reviewing equivocal cases together. Regression analyses were performed using http://vassarstats.net/multU.html

## RESULTS

The cohort of 100 gliomas with NGS and TMB data included 70 grade IV GBMs, 13 grade III astrocytomas, 4 grade II astrocytomas, 2 grade III oligodendrogliomas, 7 grade II oligodendrogliomas, and 4 grade I gliomas (**Table 1**). Eight (8%) showed loss of at least one MMR protein by IHC (**Figure 1, Table 2**). All 8 gliomas with MMR loss had TMB >20, and all 8 had mutations and/or copy number losses in corresponding MMR genes (**Table 2**). Of the remaining 92 gliomas with intact MMR IHC, only one was hypermutated (TMB=29.5). That glioma did not have mutations in *MSH2, MSH6, MLH1*, or *PMS2*, but instead contained an inactivating splice site mutation in *ATM* (**Table 2**). Sensitivity, specificity, and overall accuracy of MMR IHC for identifying hypermutated gliomas in this cohort was 88%, 100%, and 99%, respectively.

**Table 2:**
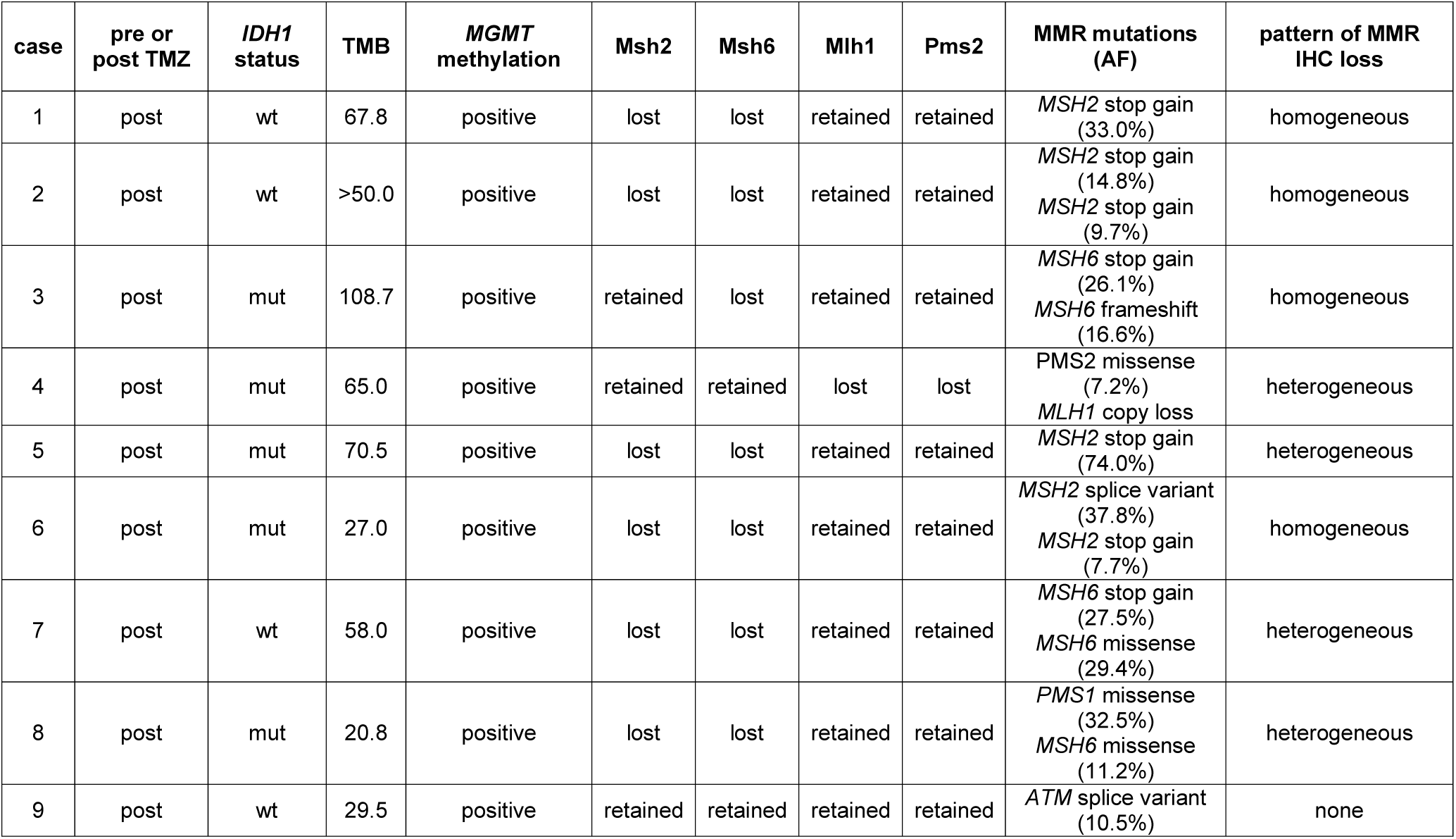
Key characteristics of all identified hypermutated gliomas. TMB=tumor mutation burden, AF=allelic fraction, IHC=immunohistochemistry.

**Figure 1:**
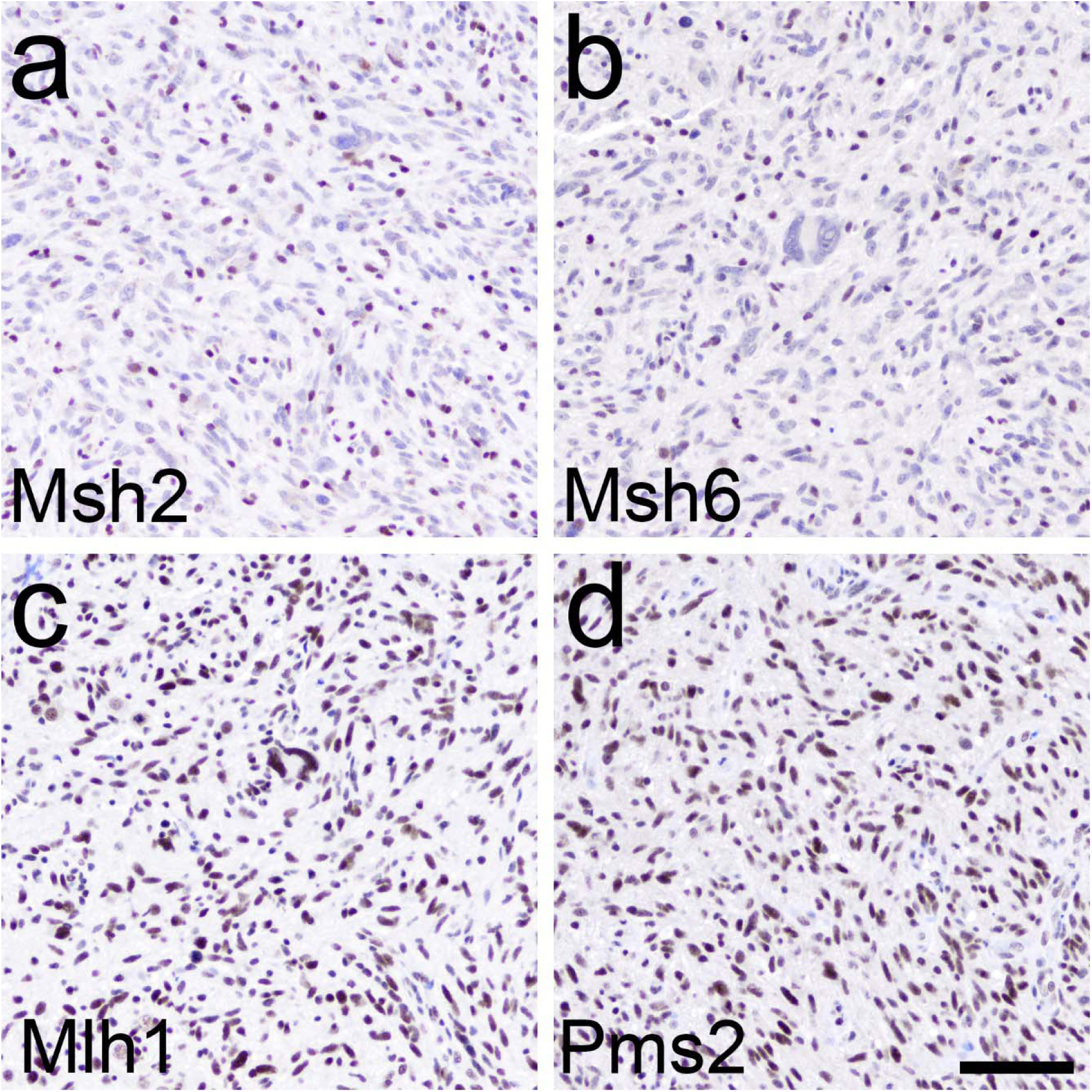
MMR IHC in Case 1. The tumor was a recurrent GBM, post-TMZ therapy, in a 57 year-old woman (**Table 2**). Tumor cells showed loss of Msh2 (a) and Msh6 (b), and retention of Mlh1 (c) and Pms2 (d). Note the normal immunostaining within nonneoplastic cells scattered throughout the tumor in (a) and (b). Scale bar=100 µm.

All 9 hypermutated gliomas had *MGMT* promoter methylation and were post-TMZ recurrences, and 5/9 (56%) were *IDH1* mutant (**Table 2**). Correlation and multiple regression analyses confirmed that the variables most strongly associated with hypermutation were prior treatment with TMZ, *MGMT* methylation, and *IDH1* mutation (**Table 3** and **Table 4**).

**Table 3:** Correlation matrix. TMZ=temozolomide.

|  | age | sex | WHO grade | astrocytoma | *IDH1* mutant | *MGMT* methylated | TMZ-treated | hypermutable |
| --- | --- | --- | --- | --- | --- | --- | --- | --- |
| age | 1 |  |  |  |  |  |  |  |
| sex | 0.053 | 1 |  |  |  |  |  |  |
| WHO grade | 0.6 | 0.084 | 1 |  |  |  |  |  |
| astrocytoma | 0.28 | 0.01 | 0.48 | 1 |  |  |  |  |
| *IDH1* mutant | -0.50 | 0.058 | -0.36 | -0.44 | 1 |  |  |  |
| *MGMT* methylated | -0.20 | -0.006 | -0.11 | -0.27 | 0.48 | 1 |  |  |
| TMZ-treated | 0.15 | 0.075 | 0.39 | 0.21 | -0.12 | 0.16 | 1 |  |
| hypermutable | 0.046 | 0.06 | 0.18 | 0.099 | 0.14 | 0.34 | 0.47 | 1 |

**Table 4:** Multiple regression results. Variables are listed from largest to smallest regression coefficients, relative to hypermutation. TMZ=temozolomide. Adjusted multiple R^2^ = 0.26 with standard error=0.24.

| variable | regression coefficient |
| --- | --- |
| TMZ-treated | 0.19 |
| *MGMT* methylated | 0.083 |
| *IDH1* mutant | 0.024 |
| astrocytoma | 0.013 |
| age | 0.0037 |
| sex | 0.001 |
| WHO grade | -0.0004 |

In 4 of 8 gliomas with lost MMR expression, the pattern of loss was clearly heterogeneous, as some tumor cells retained all MMR enzymes, while other cells lost expression of one or more MMR enzymes (**Figure 2**).

**Figure 2:**
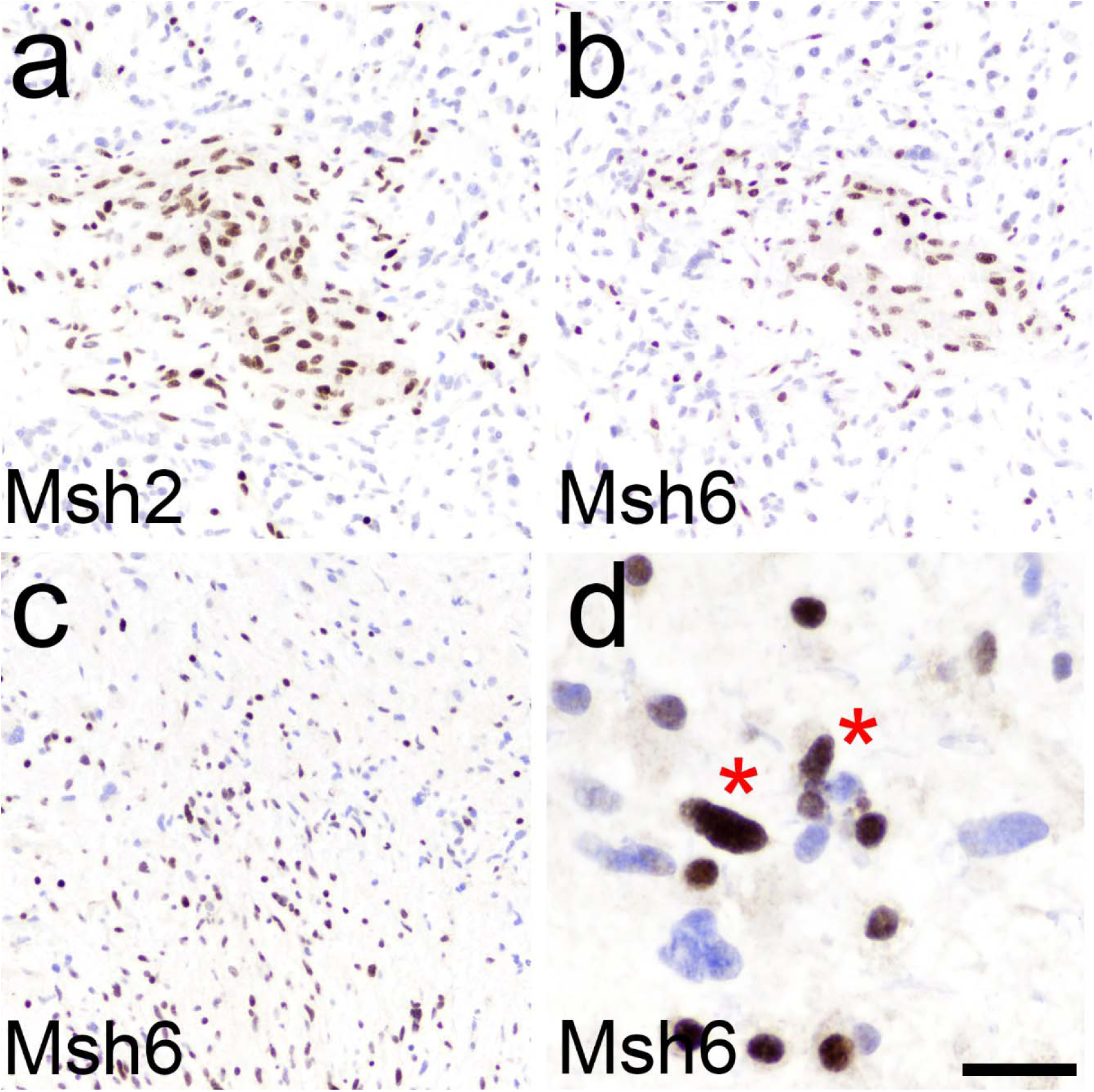
Heterogeneous MMR IHC in hypermutated gliomas. In Case 5, which was a post-TMZ *IDH1* mutant GBM in a 44 year-old woman (**Table 2**), clusters of tumor cells retained Msh2 and Msh6 positivity, but were surrounded by Msh2/6-deficient cells (a, b). Case 7 was a post-TMZ *IDH1* wild-type GBM in a 65 year-old woman (**Table 2**). Msh6 was lost in many glioma cells (c), but under high power, it was apparent that a subset of cells with identical tumor nuclear morphology retained Msh6 (d, red asterisk). Also note the smaller rounded nuclei in (d), which are most likely lymphocytes and/or oligodendrocytes. Scale bar=100 µm in (a, b, c) and 25 µm in (d).

## DISCUSSION

Despite their generally aggressive behavior, gliomas tend to have low TMB relative to most other kinds of cancer [2]. However, gliomas that are hypermutated, either at initial presentation or recurrence, may be ideal targets for immunotherapy. Such gliomas usually show increased numbers of infiltrating CD8+ cytotoxic T cells [17, 27], which is consistent with the postulate that hypermutated gliomas are more immunogenic.

Hypermutated gliomas have been a subject of intense investigation for some time, though the reported frequencies of hypermutation vary markedly due to differences in cohort selection. In our screening of over 660 untreated sporadic grade II-IV TCGA gliomas in GlioVis [5], only 15 had detectable mutations in DNA repair enzymes (not shown). But a study by the TCGA consortium showed that 7/19 (36%) TMZ-treated GBMs were hypermutated [1]. Johnson et al. reported hypermutation patterns in 6/10 (60%) post-TMZ tumors, and suggested that most of the acquired mutations were likely directly induced by TMZ [14]. In a separate study of 114 matched pre- and post-treatment GBMs, 17 (15%) showed a hypermutator profile at recurrence; among those 17 cases, 16 had mutations in MMR genes, and showed enrichment for *MGMT* methylation and *IDH1* mutation [26]. Others have verified the association between *IDH1* mutations and hypermutation after TMZ [14, 25, 30]. Among 157 pediatric gliomas, only 9 (6%) were hypermutated, and 7 contained mutations in DNA repair genes [13]. In our own cohort, 9/100 gliomas were hypermutated, all 9 had been previously treated with TMZ, all 9 had *MGMT* promoter methylation, and 5/9 were *IDH1* mutant (**Table 2**). Screening for hypermutation-associated MMR defects therefore appears to be of greatest value in recurrent, post-TMZ gliomas, especially *MGMT*-methylated and/or *IDH1* mutant tumors (**Table 3** and **Table 4**).

Although the Msh2, Msh6, Mlh1, and Pms2 IHC panel is used to screen colorectal cancer, mutations in other DNA repair genes have also been reported in post-TMZ hypermutated gliomas, including *MSH4, MSH5, MLH3, PMS1, POLE*, and *POLD1* [5, 10, 13, 26]. In our own cohort, we found a hypermutated glioma with an inactivating mutation in yet another gene associated with DNA repair, *ATM* (**Table 2**) [3]. Thus, while the MMR IHC panel designed for colorectal cancer detects most hypermutated gliomas, it may be helpful to add more IHC markers for gliomas.

Interpretation of MMR IHC in gliomas is relatively straightforward, especially since nonneoplastic cells within the glioma serve as a reliable positive control (**Figure 1**). MMR staining is lost in areas of necrosis and thermal artifact (not shown), so such regions should not be scored.

Thus far, results from immune checkpoint inhibitors in gliomas have been mixed [6, 15, 21, 29]. While at least partial responses to immune checkpoint inhibitors have been observed in patients whose sporadic gliomas had elevated TMB [8, 16], the best responses have mostly been in glioma patients with an inherited defect in an MMR gene, where 100% of the glioma cells have MMR deficiency [4, 12, 23]. Our data showing frequent heterogeneity of MMR loss in hypermutated gliomas (**Figure 2** and **Table 2**) underscores the fact that TMB is just a mathematical average of the specimen that was submitted for NGS, and that subclones may exist in “hypermutated” gliomas that are not necessarily hypermutated. Conversely, hypermutated subclones could potentially exist in tumors whose overall TMB has not yet reached the widely accepted cutoff of 20 mutations/Mb, although we did not see this in our own cohort (not shown).

In sum, DNA MMR enzyme IHC can serve as a rapid, low-cost method of screening for hypermutated gliomas. Highest yield for screening includes recurrent gliomas with *MGMT* promoter methylation and/or *IDH1* mutations. While the current panel used for colorectal cancers has very good sensitivity and excellent specificity, adding more DNA repair markers would further enhance its value. Finally, it may be valuable to consider heterogeneity in hypermutated gliomas as a possible predictor of immunotherapy response.

## Data Availability

No genomic or other publicly available datasets exist in this manuscript.

## Data Availability

No genomic or other publicly available datasets exist in this manuscript.

## AUTHOR CONTRIBUTIONS

CH and MM designed the project, performed data collection, and scored all cases. KM, RJ, and KK collected the cases, and AS performed MMR IHC. MM and CH wrote the manuscript.

## ACKNOWLEDGMENTS

This work was supported by National Institutes of Health grant R01NS102669 (C.H.) and by the Northwestern SPORE in Brain Cancer P50CA221747. MM was supported by an institutional grant from the Northwestern University Department of Pathology Resident Research Committee. The authors thank Kimberley Liggett and Juan Cuellar for their assistance with slide retrieval.

